# Protection Across Age Groups of BNT162b2 Vaccine Booster against Covid-19

**DOI:** 10.1101/2021.10.07.21264626

**Authors:** Yinon M. Bar-On, Yair Goldberg, Micha Mandel, Omri Bodenheimer, Laurence Freedman, Sharon Alroy-Preis, Nachman Ash, Amit Huppert, Ron Milo

## Abstract

**BACKGROUND:** Following administration to persons 60+ years of age, the booster vaccination campaign in Israel was gradually expanded to younger age groups who received a second dose >5 months earlier. We study the booster effect on COVID-19 outcomes.

**METHODS:** We extracted data for the period July 30, 2021 to October 6, 2021 from the Israeli Ministry of Health database regarding 4,621,836 persons. We compared confirmed Covid-19 infections, severe illness, and death of those who received a booster ≥12 days earlier (booster group) with a nonbooster group. In a secondary analysis, we compared the rates 3-7 days with ≥12 days after receiving the booster dose. We used Poisson regressions to estimate rate ratios after adjusting for possible confounding factors.

**RESULTS:** Confirmed infection rates were ≈10-fold lower in the booster versus nonbooster group (ranging 8.8-17.6 across five age groups) and 4.8-11.2 fold lower in the secondary analysis. Severe illness rates in the primary and secondary analysis were 18.7-fold (95% CI, 15.7-22.4) and 6.5-fold (95% CI, 5.1-8.3) lower for ages 60+, and 22.0-fold (95% CI, 10.3-47.0) and 3.2-fold (95% CI, 1.1-9.6) lower for ages 40-60. For ages 60+, COVID-19 associated death rates were 14.7-fold (95% CI, 9.4-23.1) lower in the primary analysis and 4.8-fold (95% CI, 2.8-8.2) lower in the secondary analysis.

**CONCLUSIONS:** Across all age groups, rates of confirmed infection and severe illness were substantially lower among those who received a booster dose of the BNT162b2 vaccine.

## Introduction

Following the resurgence of confirmed infections and severe illness in Israel ^1^, Israeli authorities approved on July 30, 2021 the administration of a BNT162b2 vaccine booster dose for persons 60 years of age or older (60+). Following promising indications that the booster dose was effective in reducing confirmed infections and severe disease against the currently dominant Delta variant for the elderly population^2^, the booster campaign was extended to younger age groups in a stepwise manner: on August 13, 2021 for ages 50-59, on August 20, for ages 40-49, on August 24, for ages 30-39, and finally, on August 29, a booster dose was approved for the entire 16+ years of age population who had received their second dose more than five months previously.

While observational studies suggest the booster dose is effective against both confirmed infection and severe disease in the elderly population, it is still not clear the extent of protection of an additional dose in younger age groups. Here, we quantified the booster effect relying on the analytical framework used to estimate the effectiveness of the booster dose for the 60+ population. The results also extend the follow-up of our previous analysis regarding the post-vaccination effect of the booster dose on the 60+ age group^2^.

## Methods

### STUDY POPULATION

Our analysis is based on medical data from the Israeli Ministry of Health database. Following Bar-On et al. ^2^, we extracted on October 6, 2021, data regarding Israeli residents 16+ years of age who had been fully vaccinated (i.e., received two doses of BNT162b2) at least five months before the end of the study and were alive on the date their age group was eligible for the booster dose, totaling 5,040,499 individuals. Similarly to Bar-On et al. ^2^, we excluded from the analysis individuals whose data did not include information regarding sex or area of residence; who had received a positive COVID-19 polymerase chain reaction (PCR) test result before the date their age group was eligible; had received a booster dose before July 30, 2021; have been abroad during the entire study period; or had been fully vaccinated before January 16, 2021. A total of 4,621,836 individuals met the inclusion criteria for the analysis (Figure 1).

**Figure 1.**
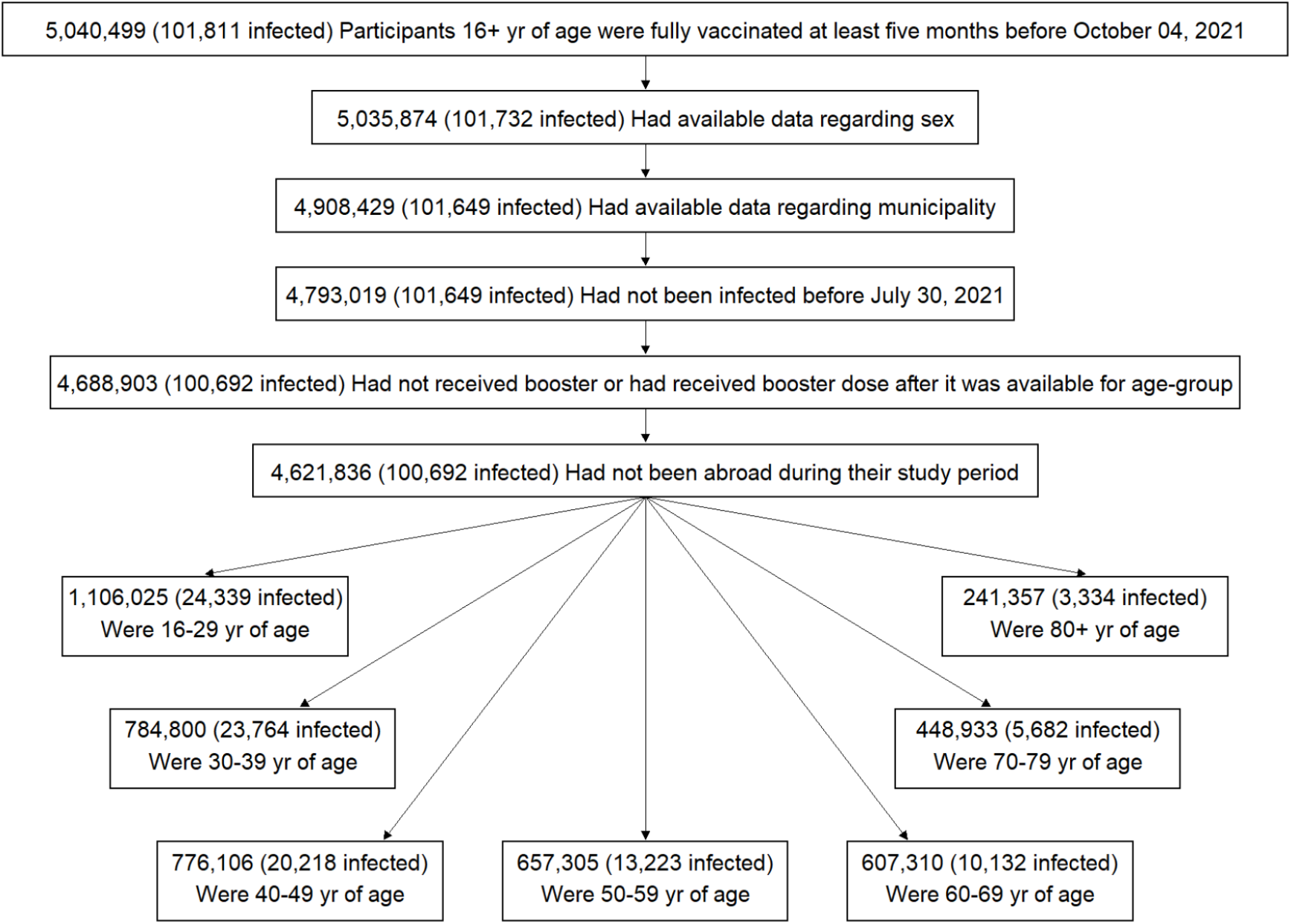
Study population.

The extracted data included vaccination dates (first, second, and third doses); information regarding PCR testing (sampling dates and results); the date of any COVID-19 related hospitalization; demographic variables, such as age, sex, area of residence, and demographic group (general Jewish, Arab, or ultra-Orthodox Jewish population), as determined by the participant’s statistical area of residence (similar to a census block)^3^; clinical status (mild or severe disease), and mortality indicator. Severe disease was defined as a resting respiratory rate of more than 30 breaths per minute, less than 94% oxygen saturation while breathing ambient air, or a ratio of partial pressure of arterial oxygen to fraction of inspired oxygen of less than 300.^4^

### STUDY DESIGN

The study period for each age group started on the date of becoming eligible to receive the booster dose. The end dates were chosen as October 4, 2021, for confirmed infection, September 29, 2021, for severe illness, and September 1, 2021 for death. The dates for severe illness and death were chosen so as to allow at least nine days for the development of severe illness and 35 days for death. We calculated the rates of confirmed infection, severe illness, and death due to COVID-19 per person-days at risk among two dynamic groups ^2^: individuals at least 12 days post booster administration (booster group) and those who had received only two vaccine doses (nonbooster group). The time of onset of severe COVID-19 was considered the test date of confirmed infection. For individuals who were abroad in part of the study period, we excluded days at risk and COVID-19 infections during their stay abroad and the 10 days following their return to Israel.

### OVERSIGHT

The study was approved by the institutional review board of the Sheba Medical Center (Helsinki approval number: SMC-8228-21). All the authors contributed to the writing and critical review of the manuscript, approved the final version, and made the decision to submit the manuscript for publication. The Israeli Ministry of Health and Pfizer have a data-sharing agreement, but only the final results of this study were shared.

### STATISTICAL ANALYSIS

We repeated the analyses conducted in Bar-On et al. (2021)^2^ with several modifications (details and comparisons with original methodology are given in Supplementary Analysis 2 in the Supplementary Appendix). Briefly, we performed a Poisson regression to estimate the rate of a specific outcome, using the function for fitting generalized linear models (glm) in R statistical software.^5^ These analyses were adjusted for the following covariates: sex, age group (16-29 years, 30-39 years, 40-49 years, 50-59 years, 60-69 years, 70-79 years, and ≥80 years), demographic group (general Jewish, Arab, ultra-Orthodox Jewish),^3^ and date of the second vaccine dose (in half-month intervals). In addition, we accounted for environmental risk by including in the model a daily exposure risk index similar to that used by Goldberg et al.^6^. Specifically, for each area of residency, we calculated the number of confirmed infections in the past seven days per 1000 residents. We included in the model 10 risk groups using the deciles of the daily exposure variable. The 7-day moving average was chosen as the number of PCR tests typically drops at weekends.

After adjusting for the above covariates, we estimated the rate ratios for confirmed infection, severe disease, and death due to COVID-19 between the booster and nonbooster groups in the different age categories by including an interaction term between age category and the booster group indicator. The age categories we considered for estimating the rate ratio were 16-29, 30-39, 40-49, 50-59, and +60 years for confirmed infection, 40-59 and 60+ years for severe disease, and 60+ for death due to COVID-19. We grouped all people aged 60 or older to compare our updated results with our previous estimates^2^ (see Supplementary Analysis 3 and Tables S11 and S12 for the results without such grouping). We grouped ages 40-49 and 50-59 when analyzing severe disease, and analyzed death due to COVID-19 only for age group 60+ due to small numbers of cases in these age groups. The average between-group rate difference ^7^ was also estimated for the different age groups. Uncertainty around the estimates was calculated by the exponent of the 95% confidence interval for the regression coefficient without adjustment for multiplicity. In a secondary analysis, we repeated the Poisson regression analysis described above but compared the rates of confirmed infection and severe COVID-19 at 3-7 days post booster administration with that at 12 days or more post booster administration.

In an additional descriptive analysis, we calculated the rate ratio of confirmed infection as a function of time from receiving the booster dose. To this end, for each age group (16-29, 30-39, 40-49, 50-59, and 60+ years), we fitted a Poisson regression that included days after the booster dose as factors in the model. The period before receipt of the booster dose was used as the reference category. As a sensitivity analysis, we analyzed the data using an alternative statistical method that relies on matching, similar to the method used by Dagan et al. ^8^ (see Supplementary Analysis 1 section in the Supplementary Appendix).

## Results

### STUDY POPULATION

Table 1 presents the characteristics of the individuals in the booster and nonbooster groups in terms of person-days at risk. We provide the same information within each age group in Tables S3-S7. The table summarizes data only for person-days used in the main analysis.

**Table 1:** Demographic and clinical characteristics of the two study cohorts. The table presents the proportion of person-days at risk instead of the proportion of individuals. Only person-days and events that were used in the main analysis are presented. Values are presented for the study period - July 30, 2021, to October 4, 2021.

| Group | nonbooster cohort<br>Person-days at risk = 90,707,958 |  |  |  | Booster cohort<br>Person-days at risk = 86,105,516 |  |  |  |
| --- | --- | --- | --- | --- | --- | --- | --- | --- |
|  | % person days at risk | # positive infections | # severe COVID-19 | # COVID-19 deaths | % person days at risk | # positive infections | # severe COVID-19 | # COVID-19 deaths |
| Female | 52.2% | 45,769 | 474 | 104 | 50.9% | 2,760 | 60 | 8 |
| Male | 47.8% | 35,131 | 666 | 174 | 49.1% | 2,948 | 98 | 15 |
| Age 16-29 | 26.4% | 21,649 | 8 | 0 | 8.2% | 267 | 0 | 0 |
| Age 30-39 | 19.6% | 20,736 | 15 | 1 | 8.4% | 758 | 1 | 0 |
| Age 40-49 | 17.0% | 16,378 | 46 | 2 | 13.3% | 1,054 | 3 | 0 |
| Age 50-59 | 13.1% | 9,912 | 114 | 5 | 16.5% | 935 | 4 | 0 |
| Age 60-69 | 12.3% | 6,881 | 227 | 38 | 23.5% | 1,197 | 29 | 4 |
| Age 70-79 | 7.3% | 3,433 | 306 | 73 | 19.8% | 867 | 41 | 7 |
| Age 80+ | 4.3% | 1,911 | 424 | 159 | 10.4% | 630 | 80 | 12 |
| General Jewish | 71.4% | 60,019 | 904 | 238 | 89.0% | 4,835 | 141 | 21 |
| Arab | 23.0% | 13,464 | 176 | 27 | 6.9% | 463 | 9 | 1 |
| Ultra-Orthodox Jewish | 5.6% | 7,417 | 60 | 13 | 4.1% | 410 | 8 | 1 |
| Vaccine Period Jan, 16-31 | 13.1% | 9,392 | 443 | 130 | 43.9% | 2,344 | 93 | 19 |
| Vaccine Period Feb, 1-15 | 16.8% | 13,459 | 368 | 95 | 29.3% | 1,676 | 46 | 2 |
| Vaccine Period Feb, 16-28 | 15.2% | 13,993 | 113 | 25 | 13.9% | 941 | 11 | 1 |
| Vaccine Period Mar, 1-15 | 21.1% | 18,636 | 107 | 21 | 9.3% | 531 | 6 | 1 |
| Vaccine Period Mar, 16-31 | 25.7% | 20,614 | 87 | 7 | 3.3% | 201 | 2 | 0 |
| Vaccine Period Apr, 1-15 | 6.6% | 4,193 | 20 | 0 | 0.3% | 15 | 0 | 0 |
| Vaccine Period Apr, 16-30 | 1.3% | 573 | 2 | 0 | 0.0% | 0 | 0 | 0 |
| Vaccine Period May, 1-15 | 0.1% | 40 | 0 | 0 | 0.0% | 0 | 0 | 0 |

The nonbooster group includes approximately 91 million person-days with 80,900 confirmed infections, 1,140 cases of severe illness, and 278 deaths. The booster group includes approximately 86 million person-days with 5,708 confirmed infections, 158 cases of severe illness, and 23 deaths. In comparison to the nonbooster group, the booster group had more men (49.1% vs. 47.8%), a higher representation of the general Jewish population (89.0% vs. 71.4%), more individuals 70+ years of age (30.2% vs. 11.6%), fewer individuals below 40 years of age (16.6% vs. 46.0%) and a greater number of individuals who received their second vaccination dose in January 2021 (43.9% vs. 13.1%). These substantial between-group differences were adjusted for by including the variables as covariates in the Poisson regression model.

### EFFECT OF THE BOOSTER DOSE ACROSS AGE GROUPS

The detailed results of the Poisson regression analysis for confirmed infection, severe illness, and death are provided in Table S8-S10 and are summarized in Table 2 and in Table 3. The rate of confirmed infection was lower in the booster group than in the nonbooster group by a similar factor across the age groups: 12.4 (95% confidence interval [CI], 11.9 to 12.9) for people 60+ years of age, 12.2 (95% CI, 11.4 to 13.1) for people aged 50-59, 9.7 (95% CI, 9.2 to 10.4) for people aged 40-49, 8.8 (95% CI, 8.2 to 9.5) for people aged 30-39, and 17.6 (95% CI, 15.6 to 19.9) for people aged 16-29. Interestingly we find a higher increase in protection at the youngest age group (16-29). The absolute between-group difference in the rate of confirmed infection was 61.8 infections per 100,000 person-days for people 60+ years of age, 75.2 for people aged 50-59, 89.4 for people aged 40-49, 97.7 for people aged 30-39, and 80.2 for people aged 16-29. In the secondary analysis, we saw a similar pattern, namely, the rate of confirmed infection after at least 12 days from receipt of the vaccine was substantially lower than the rate 3 to 7 days after booster receipt: 7.4 (95% CI, 7.0 to 7.8) for people 60+ years of age, 7.3 (95% CI, 6.7 to 7.9) for people aged 50-59, 5.4 (95% CI, 5.0 to 5.8) for people aged 40-49, 4.8 (95% CI, 4.4 to 5.2) for people aged 30-39, and 11.2 (95% CI, 9.9 to 12.8) for people aged 16-29.

**Table 2.** Summary of the results regarding confirmed infections of the Poisson regression analysis for the booster group and nonbooster group, for the different age groups. For each group, we provide the number of confirmed infections, the total number of person-days at risk, and the estimated rate ratio for the primary analysis (nonbooster relative to at least 12 days post booster-vaccination) and the secondary analysis (3-7 days post booster-vaccination relative to at least 12 days post booster-vaccination).

| Age | Nonbooster group infections<br>(person-days at risk) | Booster group infections - day 12+<br>(person-days at risk) | Booster control group infections - day 3-7<br>(person-days at risk) | Rate ratio day 12+ relative to nonbooster<br>[95% CI] | Rate ratio day 12+ relative to day 3-7<br>[95% CI] |
| --- | --- | --- | --- | --- | --- |
| 60+ | 12,225<br>(21,660,770) | 2,694<br>(46,201,515) | 2,395<br>(5,628,282) | 12.4<br>[11.9, 12.9] | 7.4<br>[7.0, 7.8] |
| 50-59 | 9,912<br>(11,887,725) | 935<br>(14,204,942) | 1,453<br>(2,433,725) | 12.2<br>[11.4, 13.1] | 7.3<br>[6.7, 7.9] |
| 40-49 | 16,378<br>(15,416,326) | 1,054<br>(11,409,730) | 1,701<br>(2,552,036) | 9.7<br>[9.2, 10.4] | 5.4<br>[5.0, 5.8] |
| 30-39 | 20,736<br>(17,757,731) | 758<br>(7,228,945) | 1,398<br>(2,085,818) | 8.8<br>[8.2, 9.5] | 4.8<br>[4.4, 5.2] |
| 16-29 | 21,649<br>(23,985,406) | 267<br>(7,060,384) | 1,503<br>(2,555,298) | 17.6<br>[15.6, 19.9] | 11.2<br>[9.9, 12.8] |

**Table 3.**
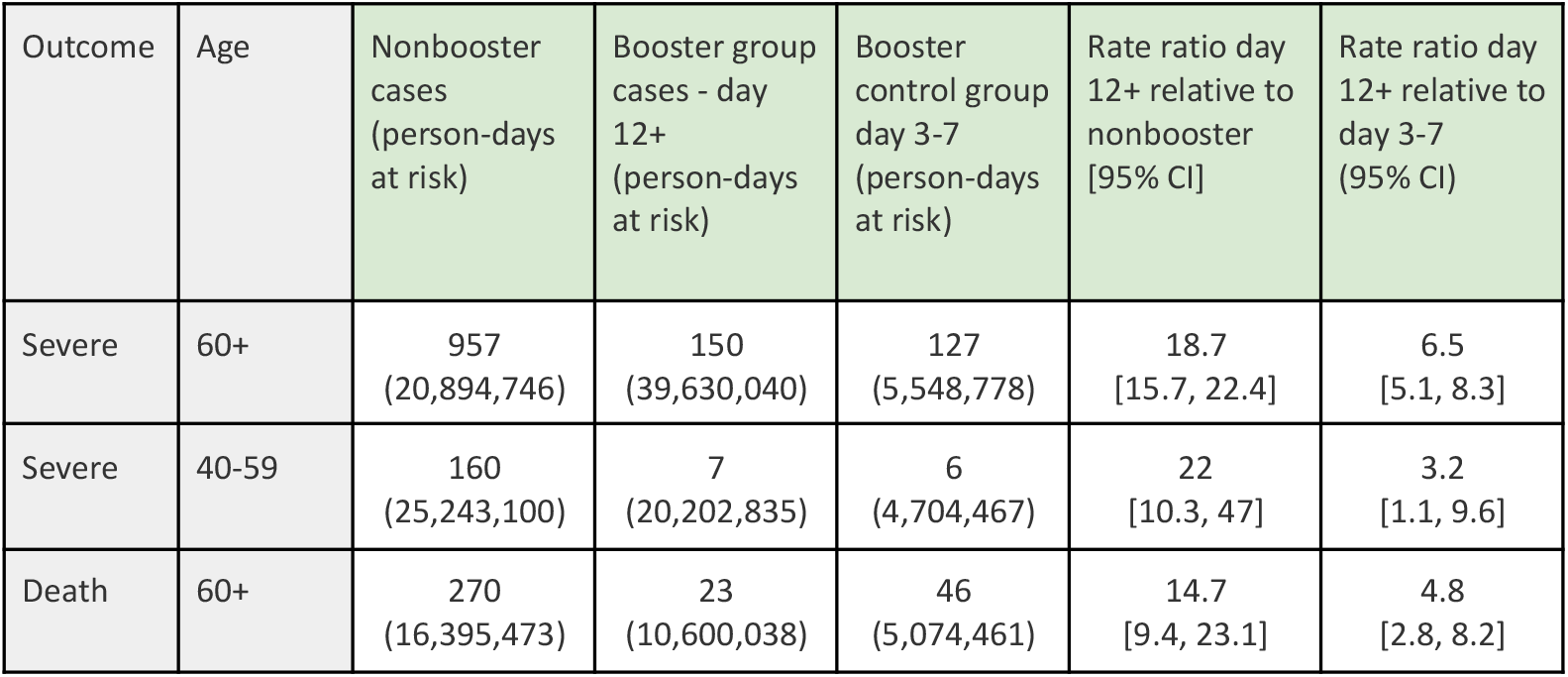
Summary of results regarding severe cases and death due to COVID-19 in the Poisson regression analysis for the booster group and nonbooster group across age groups. For each outcome, we provide the number of COVID-19 cases, the total number of person-days at risk and the estimated rate ratio for the primary analysis (nonbooster relative to at least 12 days post-vaccination) and the secondary analysis (3-7 days post-vaccination relative to at least 12 days post-vaccination).

| Outcome | Age | Nonbooster cases (person-days at risk) | Booster group cases - day 12+ (person-days at risk) | Booster control group day 3-7 (person-days at risk) | Rate ratio day 12+ relative to nonbooster [95% CI] | Rate ratio day 12+ relative to day 3-7 (95% CI) |
| --- | --- | --- | --- | --- | --- | --- |
| Severe | 60+ | 957<br>(20,894,746) | 150<br>(39,630,040) | 127<br>(5,548,778) | 18.7<br>[15.7, 22.4] | 6.5<br>[5.1, 8.3] |
| Severe | 40-59 | 160<br>(25,243,100) | 7<br>(20,202,835) | 6<br>(4,704,467) | 22<br>[10.3, 47] | 3.2<br>[1.1, 9.6] |
| Death | 60+ | 270<br>(16,395,473) | 23<br>(10,600,038) | 46<br>(5,074,461) | 14.7<br>[9.4, 23.1] | 4.8<br>[2.8, 8.2] |

The rate of severe illness was lower in the booster group than in the nonbooster group across the two age groups studied: 18.7 (95% CI, 15.7 to 22.4) for people 60+ years of age, and 22.0 (95% CI, 10.3 to 47.0) for people aged 40-59. The absolute difference in the rate of severe illness between the booster and non booster group was 5.9 cases per 100,000 person-days for people aged 60 years or older, and an order of magnitude lower, 0.66 for people aged 40-59. In the secondary analysis, the rate of severe illness 12+ days after the booster was lower than the rate 3 to 7 days after the booster by a factor of 6.5 (95% CI, 5.1 to 8.3) in the 60+ years of age group, and by a factor of 3.2 (95% CI, 1.1 to 9.6) in the 40-69 age group. The rate of severe disease in the youngest age groups is very low and there were not enough cases to compare the rates of severe disease (23 in the nonbooster group and one in the 12+ days post booster and in the booster control groups).

Due to small numbers, the analysis for death due to COVID-19 was performed only for people ages 60 and above. For this outcome, the rate in the booster group was lower than in the nonbooster group by a factor of 14.7 (95% CI, 9.4 to 23.1). The absolute difference between the booster and nonbooster group was 2.1 cases per 100,000 person-days. In the secondary analysis, the rate of severe illness 12+ days after the booster was lower than the rate 3 to 7 days after the booster by a factor of 4.8 (95% CI, 2.8 to 8.2).

We also estimated the reduction in the rate of confirmed infection in the booster group compared with the nonbooster group as a function of time from booster vaccination across the different age groups. As shown in Figure 2, the different age groups showed very similar dynamics, where from 12 days onward, the rate of confirmed infection was about 10-fold lower compared with the nonbooster group, suggesting the effect of the booster dose was retained throughout the study period. Immediately after vaccination, the rate of confirmed infection in the booster cohort was lower than that in the nonbooster group in all age groups, a difference that is probably the result of behavioral changes that often follow vaccination as detailed in the discussion.

**Figure 2.**
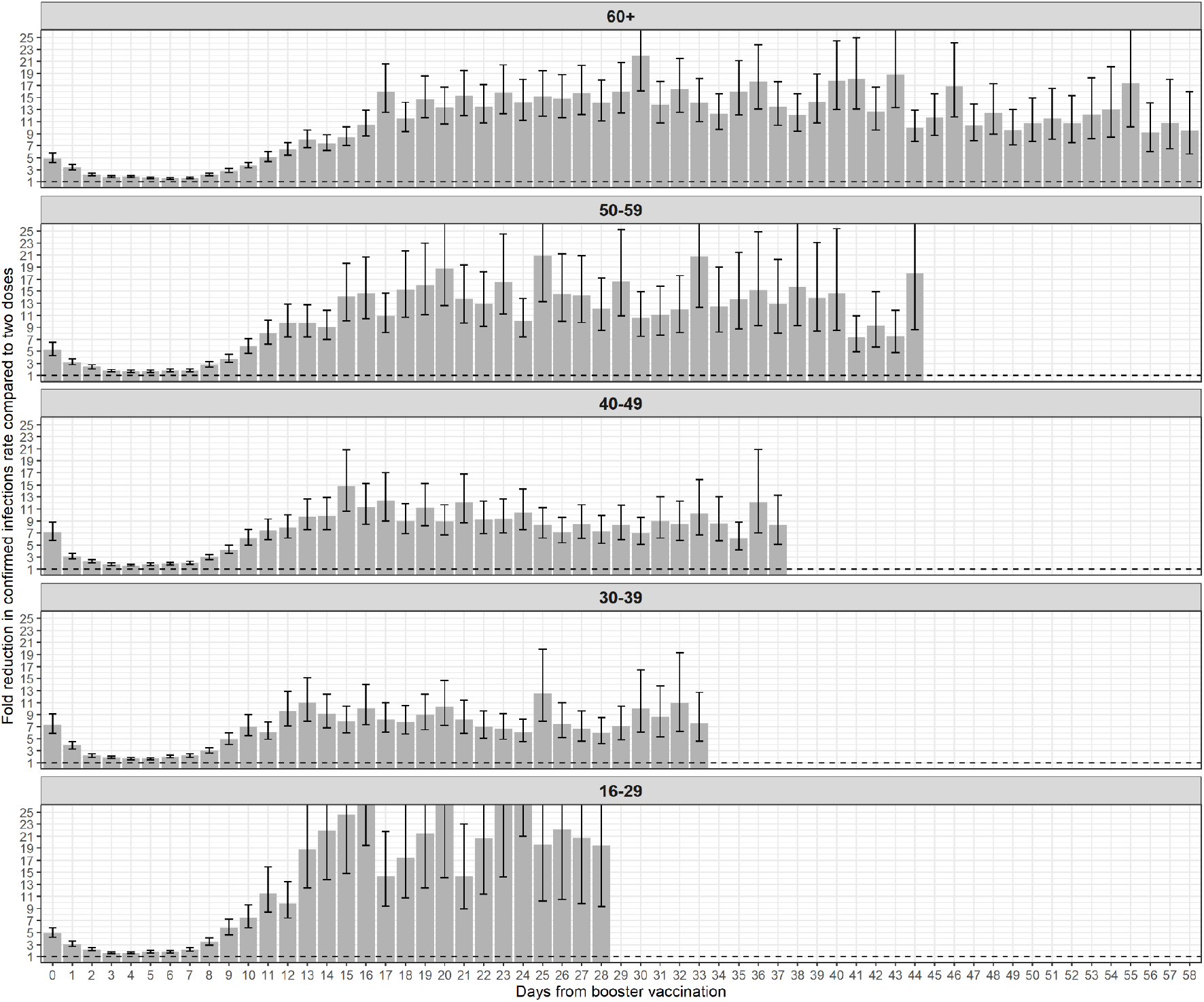
The fold reduction in the rate of confirmed infections in the booster group compared to the nonbooster group as a function of the number of days following the booster dose (day 0 = day receiving the booster dose), for the different age groups. The dashed line represents no added protection by the booster dose. Error bars are 95% confidence intervals not corrected for multiplicity.

We performed a sensitivity analysis that uses matching resulted in the following estimates for the rate ratio of confirmed infection between the booster and nonbooster groups: 9.7 (95% CI, 7.6 to 12.8) for persons aged 60 or older, 10.0 (95% CI, 7.9 to 12.6) for persons aged 50-59, 8.6 (95% CI, 6.8 to 10.2) for persons aged 40-49, 7.7 (95% CI, 5.3 to 9.4) for persons aged 30-39, 16.4 (95% CI, 11.8 to 22.1) for persons aged 16-29. For severe illness, this approach yielded an estimated rate ratio of 13.1 (95% CI, 7.0 to 33.7) for persons aged 60 years or above.

## Discussion

Our study seeks to determine whether the booster dose had a similar effect in different age groups. We demonstrate that the booster dose reduces the rate of confirmed infection and severe illness by a similar factor in the age groups studied (for the youngest age group a higher increase in the rate ratio was observed). The dynamics of the rate ratio between the booster and nonbooster groups show a similar pattern across the age groups. These findings are consistent with those of the phase 2/3 clinical trial of the BNT162b2 vaccine ^9^, where vaccine efficacy was similar across age groups.

In addition to demonstrating the effect of the booster dose across age groups, we extend the follow-up period of our previous analysis regarding the effect of the booster dose in people 60+ years of age^2^ (45 days instead of 25 days). As shown in Figure 2, the effect of the booster dose remains stable throughout the two months observation period.

Although our analysis attempts to address confounding and detection bias, some sources of bias may not have been measured or corrected adequately. These biases might include differences between the population of booster recipients and those who did not receive the booster with respect to care-seeking behaviors and/or cautiousness, along with differences in coexisting comorbidities which are not recorded in the national database. One approach to reduce the extent of confounding between the booster and nonbooster groups is by focusing on persons who received the booster dose and comparing the rates during a period in which the booster effect was expected to be small and those related to a period in which the booster had become effective. We, therefore, in our secondary analysis, compare the rates obtained for the period 12+ days from receipt of the booster with the equivalent ones for the period 3-7 from booster administration, when the booster effect is expected to be small and behavioral changes after vaccination are less marked.

While this type of analysis reduces confounding, estimates of the rate ratio during the first days after vaccination could include the effect of transient biases and thus lead to an underestimation of the effect of the booster dose. These potential biases include the “healthy-vaccinee” bias ^10^, in which people who feel ill tend not to get vaccinated in the following days, increasing the number of events in the nonbooster group in the first days following vaccination. Another source of underestimate of vaccine protection in the secondary analysis can arise from the tendency to decrease social interactions in the days of recuperation immediately after the vaccination, which would translate into less confirmed infections a few days later and a seeming protection. This secondary analysis yielded a smaller effect of about five-fold decrease in the rate of confirmed infection, observed consistently across age groups. In respect to severe illness, as presented in Table 3, the secondary analysis shows elevated protection in both age groups, but with wider confidence intervals due to the lower number of severe cases in the younger age group and especially so in the secondary booster control group which has a relatively small number of risk-days.

Compared to our previous analysis of the elderly population^2^, we made two methodological modifications in the current analysis. First, instead of including indicators for calendar dates in order to adjust for exposure risk, we calculated a spatial-temporal index of risk according to the number of infections in each town. This better measures the exposure risk of each individual, and has an additional advantage of counting more days at risk for the nonbooster group, as days can be counted starting at the date of eligibility to receive the booster dose. A second modification was in comparing the rates in the booster group to days 3-7 after receiving the booster dose instead of days 4-6 in the secondary analysis. This change was done in order to increase the number of person days at risk in this group, which enabled applying the secondary analysis also to severe COVID-19.

Understanding the protective effect of the booster dose in younger age groups is key for public health policy making and can be a way to control transmission without costly social distance measures and quarantines. Our findings provide evidence for the effectiveness of the booster dose against the currently dominant Delta variant in people 16+ years of age, as well as evidence for the maintenance of the effectiveness over an extended follow-up time for people 60+ years of age. Future studies will help determine the longer-term effectiveness of the booster dose against current and emerging variants.

## Data Availability

Aggregated data are given in the supplementary information. Personal data cannot be shared due to privacy.

## Ethics statement

The study was approved by the Institutional Review Board of the Sheba Medical Center. Helsinki approval number: SMC-8228-21.

## Competing interests statement

All authors declare no competing interests.

## Acknowledgements

We thank Ronen Fluss for productive feedback on this manuscript.

## Supplementary Appendix

### Supplementary Analysis 1 - matching based approach

In order to validate our findings, we conducted an independent analysis which relied on matching fully vaccinated individuals who received a booster dose with similar individuals who received only two vaccine doses. The approach was similar to that conducted by Dagan et.al.^8^, and aimed at comparing individuals’ risk rather than rate based on person-days. Briefly, each individual in the booster group was matched to an individual who was in the nonbooster group on the booster-vaccination day. Matching was conducted based on the following characteristics: age group (16-29, 30-39, 40-49, 50-59, 60-69, 70-79 and 80+), gender, week of second vaccine dose and demographic group (general Jewish, Arab, ultra-Orthodox). Follow-up for both individuals ended at the time of infection. Both individuals in a pair were censored at the end of the study or at the time the nonbooster individual got a booster dose. We calculated the probability of being free of infection in the two groups as a function of time using the Kaplan-Meier estimator, and compared the survival probabilities of the two groups at the end of the study. For each group, we calculated the probability of an event occurring between day 12 following the booster and the end of the study, among individuals still at risk on day 12. We used the ratio between the probabilities of the two groups as an estimate for the risk ratio for our population over the study time. We generated 95% confidence intervals around this estimate using the percentile bootstrap method with 200 repetitions.

Using this approach, we obtained the following estimates for the rate ratio of confirmed infection between the booster and nonbooster groups: 9.7 (95% CI, 7.6 to 12.8) for persons aged 60 or older, 10.0 (95% CI, 7.9 to 12.6) for persons aged 50-59, 8.6 (95% CI, 6.8 to 10.2) for persons aged 40-49, 7.7 (95% CI, 5.3-9.4) for persons aged 30-39, 16.4 (95% CI, 11.8 to 22.1) for persons aged 16-29. For severe illness, this approach yielded an estimated rate ratio of 13.1 (95% CI, 7.0 to 33.7) for persons aged 60 years or above.

### Supplementary Analysis 2 - comparison with previous analysis methods

Some of the details of our current analysis are different from our previous analysis for the 60 years or older population^2^. Specifically, in order to increase the number of person-days at risk for the control group in the secondary analysis, we changed the analysis in the following manner. Instead of using the rate of confirmed infection and severe illness 4 to 6 days post vaccination as the control group in the secondary analysis, we now use days 3 to 7. Additionally, we previously filtered out days at risk that occurred prior to 12 days after the start of the booster vaccination campaign. This was done to ensure that we collect data for the booster group as well as the control group on similar calendar days, so we could use calendar day as a covariate in the Poisson regression analysis. In the current analysis, we relaxed this filtering criterion. In order to still be able to adjust for the exposure people experience at different calendar days, we now use the weekly average incidence rate in the place of residence of each person, binned into 10 quantiles as a covariate in our analysis.

To assess the impact of these different modelling choices on the results of our analysis, we repeated the analysis using the exact methodology used in Bar-On et al.^2^. The results of this analysis are given in Tables S1 and S2, and are very similar to the results of our current analysis. As can be seen when comparing Table 3 and Table S2, the main difference between the two modelling choices is that the control group in the secondary analysis now has much more person-days contributing to it, which reduces the uncertainty range of rate ratios.

### Supplementary Analysis 3 - breakdown of protection in 60+ population

To look more closely at the effect of the booster dose among people aged 60 years or older, we repeated the same primary and secondary analysis reported in the Methods section of the main text, but using the following age groups instead of the “60+” age group: 60-69 years of age, 70-79 years of age, and people aged 80 year or older. The results of this analysis are provided in Tables S11 and S12.

### Supplementary Tables

**Table S1.** Summary of the results regarding confirmed infections of the Poisson regression analysis using the methodology in Bar-On et al. ^2^

| Age | Nonbooster group infections (person-days at risk) | Booster group infections - day 12+ (person-days at risk) | Booster control group infections - day 4-6 (person-days at risk) | Rate ratio day 12+ relative to nonbooster [95% CI] | Rate ratio day 12+ relative to day 4-6 (95% CI) |
| --- | --- | --- | --- | --- | --- |
| 60+ | 7,918<br>(11,416,889) | 2,694<br>(46,277,894) | 1,098<br>(2,430,916) | 12.5 [12, 13.1] | 7.8 [7.3, 8.4] |
| 50-59 | 5,609<br>(7,799,080) | 937<br>(14,424,641) | 442<br>(781,518) | 12.3 [11.5, 13.2] | 8.9 [7.9, 9.9] |
| 40-49 | 9,469<br>(10,280,089) | 1,056<br>(11,766,037) | 484<br>(839,249) | 11.4 [10.7, 12.2] | 6.6 [6, 7.4] |
| 30-39 | 12,377<br>(12,151,084) | 759<br>(7,519,102) | 483<br>(789,522) | 11.2 [10.4, 12] | 6.3 [5.7, 7.1] |
| 16-29 | 10,105<br>(15,184,595) | 267<br>(7,131,043) | 334<br>(827,046) | 19.5 [17.3, 22] | 11.4 [9.7, 13.3] |

**Table S2-.**
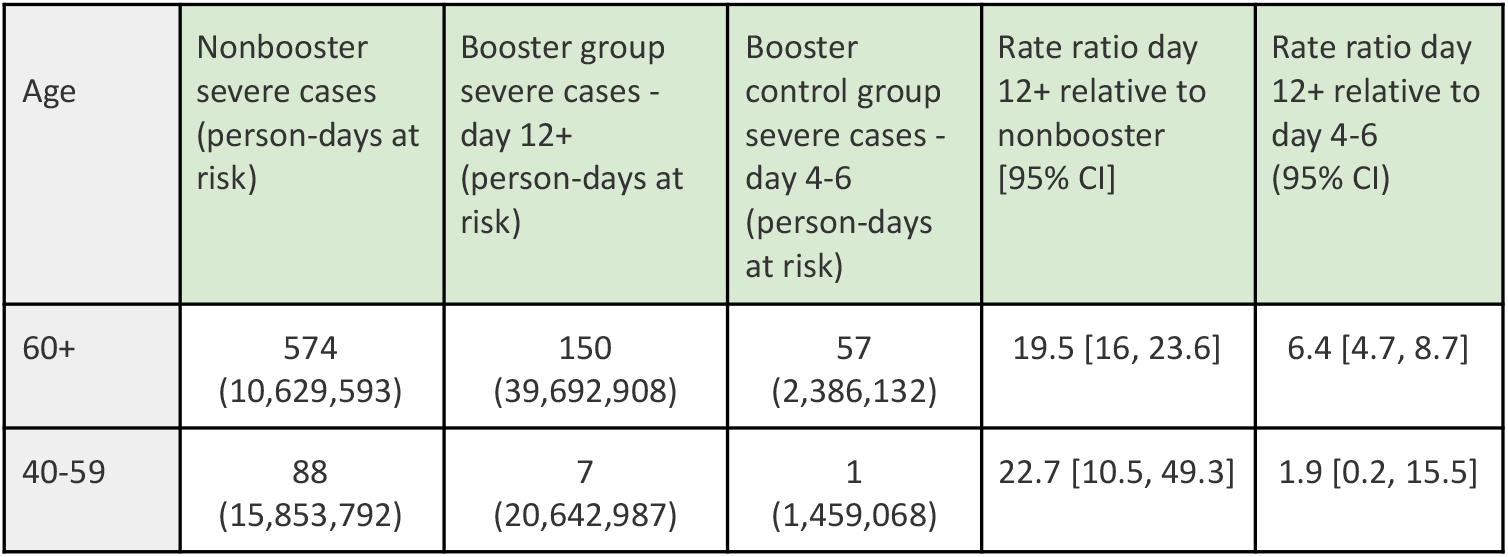
Summary of the results regarding severe COVID-19 cases of the Poisson regression analysis using the methodology in Bar-On et al. ^2^

**Table S3:** Demographic and clinical characteristics of the two study cohorts for people aged 60 or above. Only person-days and events that were used in the main analysis are presented. Values are presented for the study period - July 30, 2021, to October 4, 2021.

| Group | Nonbooster cohort<br>Person-days at risk = 21,660,770 |  |  |  | Booster cohort<br>Person-days at risk = 46,201,515 |  |  |  |
| --- | --- | --- | --- | --- | --- | --- | --- | --- |
|  | % person days at risk | # positive infections | # severe COVID-19 | # COVID-19 deaths | % person days at risk | # positive infections | # severe COVID-19 | # COVID-19 deaths |
| Female | 57.3% | 6,717 | 404 | 102 | 52.7% | 1,234 | 57 | 8 |
| Male | 42.7% | 5,508 | 553 | 168 | 47.3% | 1,460 | 93 | 15 |
| Age 60-69 | 51.4% | 6,881 | 227 | 38 | 43.8% | 1,197 | 29 | 4 |
| Age 70-79 | 30.5% | 3,433 | 306 | 73 | 36.8% | 867 | 41 | 7 |
| Age 80+ | 18.1% | 1,911 | 424 | 159 | 19.4% | 630 | 80 | 12 |
| General Jewish | 81.3% | 10,082 | 790 | 230 | 90.5% | 2,376 | 134 | 21 |
| Arab | 14.2% | 1,299 | 119 | 27 | 5.7% | 140 | 9 | 1 |
| Ultra-Orthodox Jewish | 4.5% | 844 | 48 | 13 | 3.8% | 178 | 7 | 1 |
| Vaccine Period Jan, 16-31 | 37.8% | 4,636 | 407 | 127 | 65.5% | 1,732 | 91 | 19 |
| Vaccine Period Feb, 1-15 | 32.5% | 3,967 | 316 | 91 | 27.4% | 761 | 44 | 2 |
| Vaccine Period Feb, 16-28 | 9.4% | 1,158 | 89 | 25 | 3.7% | 110 | 8 | 1 |
| Vaccine Period Mar, 1-15 | 9.1% | 1,091 | 68 | 20 | 2.4% | 66 | 5 | 1 |
| Vaccine Period Mar, 16-31 | 8.5% | 1,099 | 59 | 7 | 0.9% | 22 | 2 | 0 |
| Vaccine Period Apr, 1-15 | 2.2% | 243 | 16 | 0 | 0.1% | 3 | 0 | 0 |
| Vaccine Period Apr, 16-30 | 0.4% | 27 | 2 | 0 | 0.0% | 0 | 0 | 0 |
| Vaccine Period May, 1-15 | 0.1% | 4 | 0 | 0 | 0.0% | 0 | 0 | 0 |

**Table S4:** Demographic and clinical characteristics of the two study cohorts for people aged 50-59. Only person-days and events that were used in the main analysis are presented. Values are presented for the study period - July 30, 2021, to October 4, 2021.

| Group | Nonbooster cohort<br>Person-days at risk = 11,887,725 |  |  |  | Booster cohort<br>Person-days at risk = 14,204,942 |  |  |  |
| --- | --- | --- | --- | --- | --- | --- | --- | --- |
|  | % person days at risk | # positive infections | # severe COVID-19 | # COVID-19 deaths | % person days at risk | # positive infections | # severe COVID-19 | # COVID-19 deaths |
| Female | 52.9% | 5,530 | 41 | 2 | 49.50% | 438 | 1 | 0 |
| Male | 47.1% | 4,382 | 73 | 3 | 50.50% | 497 | 3 | 0 |
| General Jewish | 70.3% | 7,640 | 74 | 5 | 87.40% | 776 | 3 | 0 |
| Arab | 25.7% | 1,568 | 35 | 0 | 9.00% | 84 | 0 | 0 |
| Ultra-Orthodox Jewish | 4.1% | 704 | 5 | 0 | 3.60% | 75 | 1 | 0 |
| Vaccine Period Jan, 16-31 | 10.1% | 1,417 | 24 | 2 | 28.70% | 292 | 1 | 0 |
| Vaccine Period Feb, 1-15 | 26.7% | 2,994 | 41 | 2 | 46.50% | 408 | 2 | 0 |
| Vaccine Period Feb, 16-28 | 19.2% | 1,898 | 11 | 0 | 15.30% | 160 | 0 | 0 |
| Vaccine Period Mar, 1-15 | 18.7% | 1,679 | 19 | 1 | 6.80% | 50 | 1 | 0 |
| Vaccine Period Mar, 16-31 | 20.1% | 1,605 | 15 | 0 | 2.70% | 23 | 0 | 0 |
| Vaccine Period Apr, 1-15 | 4.4% | 275 | 4 | 0 | 0.20% | 2 | 0 | 0 |
| Vaccine Period Apr, 16-30 | 0.8% | 41 | 0 | 0 | 0.00% | 0 | 0 | 0 |
| Vaccine Period May, 1-15 | 0.1% | 3 | 0 | 0 | 0.00% | 0 | 0 | 0 |

**Table S5:** Demographic and clinical characteristics of the two study cohorts for people aged 49-49. Only person-days and events that were used in the main analysis are presented. Values are presented for the study period - July 30, 2021, to October 4, 2021.

| Group | Nonbooster cohort<br>Person-days at risk = 15,416,326 |  |  |  | Booster cohort<br>Person-days at risk = 11,409,730 |  |  |  |
| --- | --- | --- | --- | --- | --- | --- | --- | --- |
|  | % person days at risk | # positive infections | # severe COVID-19 | # COVID-19 deaths | % person days at risk | # positive infections | # severe COVID-19 | # COVID-19 deaths |
| Female | 51.1% | 9,050 | 18 | 0 | 49.4% | 520 | 2 | 0 |
| Male | 48.9% | 7,328 | 28 | 2 | 50.6% | 534 | 1 | 0 |
| General Jewish | 71.9% | 12,038 | 28 | 2 | 87.6% | 868 | 3 | 0 |
| Arab | 23.7% | 3,008 | 14 | 0 | 8.4% | 119 | 0 | 0 |
| Ultra-Orthodox Jewish | 4.4% | 1,332 | 4 | 0 | 4.0% | 67 | 0 | 0 |
| Vaccine Period Jan, 16-31 | 6.6% | 1,387 | 9 | 0 | 17.6% | 197 | 1 | 0 |
| Vaccine Period Feb, 1-15 | 14.3% | 2,766 | 6 | 2 | 31.4% | 315 | 0 | 0 |
| Vaccine Period Feb, 16-28 | 24.3% | 4,127 | 11 | 0 | 34.3% | 378 | 2 | 0 |
| Vaccine Period Mar, 1-15 | 22.1% | 3,531 | 13 | 0 | 12.0% | 116 | 0 | 0 |
| Vaccine Period Mar, 16-31 | 25.8% | 3,790 | 7 | 0 | 4.5% | 46 | 0 | 0 |
| Vaccine Period Apr, 1-15 | 5.8% | 682 | 0 | 0 | 0.3% | 2 | 0 | 0 |
| Vaccine Period Apr, 16-30 | 1.0% | 91 | 0 | 0 | 0.0% | 0 | 0 | 0 |
| Vaccine Period May, 1-15 | 0.1% | 4 | 0 | 0 | 0.0% | 0 | 0 | 0 |

**Table S6:**
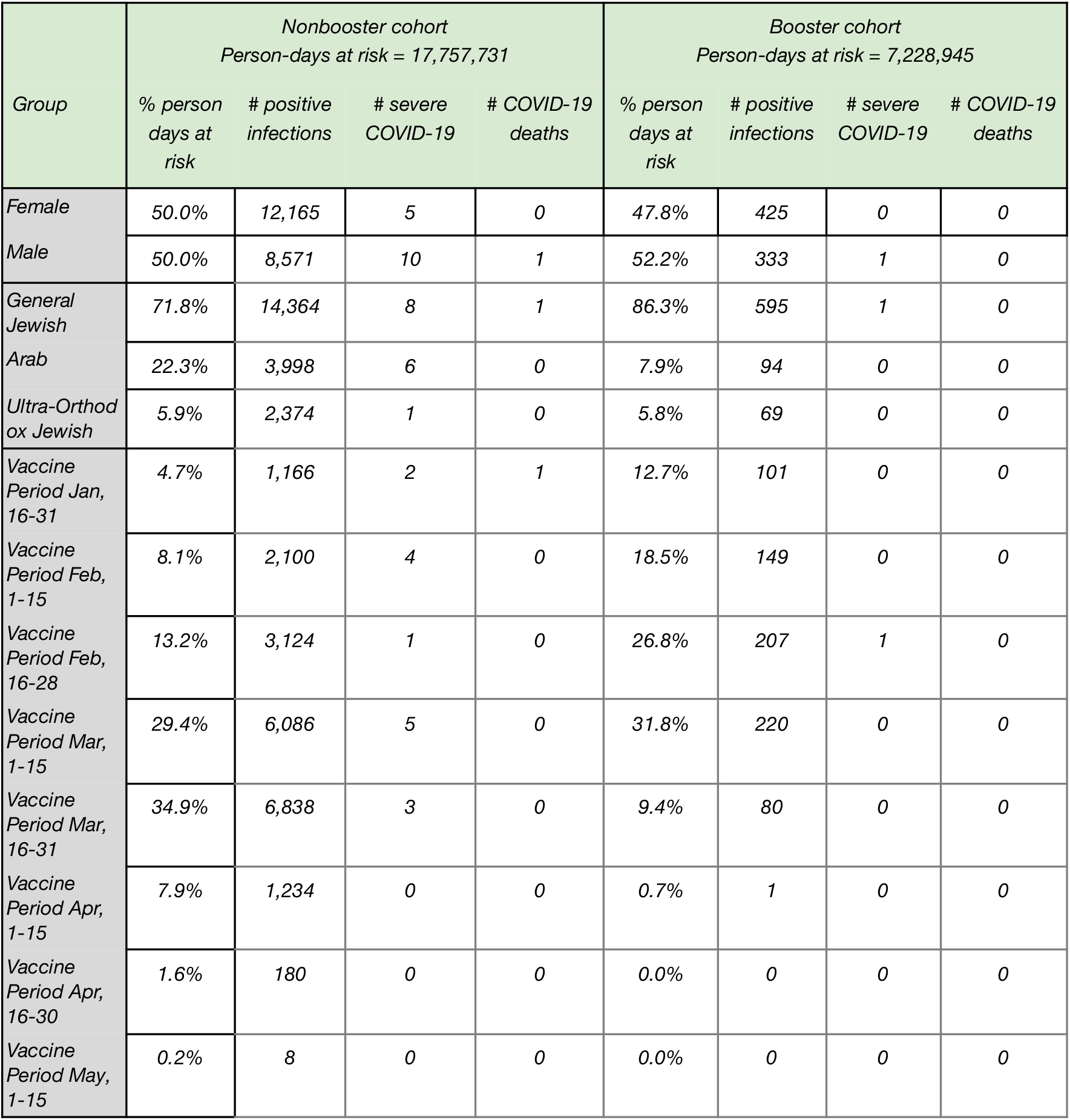
Demographic and clinical characteristics of the two study cohorts for people aged 30-39. Only person-days and events that were used in the main analysis are presented. Values are presented for the study period - July 30, 2021, to October 4, 2021.

**Table S7:** Demographic and clinical characteristics of the two study cohorts for people aged 16-29. Only person-days and events that were used in the main analysis are presented. Values are presented for the study period - July 30, 2021, to October 4, 2021.

| Group | Nonbooster cohort<br>Person-days at risk = 23,985,406 |  |  |  | Booster cohort<br>Person-days at risk = 7,060,384 |  |  |  |
| --- | --- | --- | --- | --- | --- | --- | --- | --- |
|  | % person days at risk | # positive infections | # severe COVID-19 | # COVID-19 deaths | % person days at risk | # positive infections | # severe COVID-19 | # COVID-19 deaths |
| Female | 49.5% | 12,307 | 6 | 0 | 47.6% | 143 | 0 | 0 |
| Male | 50.5% | 9,342 | 2 | 0 | 52.4% | 124 | 0 | 0 |
| General Jewish | 62.4% | 15,895 | 4 | 0 | 87.0% | 220 | 0 | 0 |
| Arab | 29.7% | 3,591 | 2 | 0 | 7.0% | 26 | 0 | 0 |
| Ultra-Orthodox Jewish | 7.9% | 2,163 | 2 | 0 | 6.0% | 21 | 0 | 0 |
| Vaccine Period Jan, 16-31 | 2.7% | 786 | 1 | 0 | 7.8% | 22 | 0 | 0 |
| Vaccine Period Feb, 1-15 | 5.7% | 1,632 | 1 | 0 | 15.1% | 43 | 0 | 0 |
| Vaccine Period Feb, 16-28 | 14.1% | 3,686 | 1 | 0 | 32.0% | 86 | 0 | 0 |
| Vaccine Period Mar, 1-15 | 26.4% | 6,249 | 2 | 0 | 32.3% | 79 | 0 | 0 |
| Vaccine Period Mar, 16-31 | 37.3% | 7,282 | 3 | 0 | 11.7% | 30 | 0 | 0 |
| Vaccine Period Apr, 1-15 | 11.3% | 1,759 | 0 | 0 | 1.2% | 7 | 0 | 0 |
| Vaccine Period Apr, 16-30 | 2.2% | 234 | 0 | 0 | 0.0% | 0 | 0 | 0 |
| Vaccine Period May, 1-15 | 0.2% | 21 | 0 | 0 | 0.0% | 0 | 0 | 0 |

**Table S8.** Poisson regression results for confirmed infection.

| *term* | *Primary analysis* |  | *Secondary analysis* |  |
| --- | --- | --- | --- | --- |
|  | *estimate* | *std.error* | *estimate* | *std.error* |
| *(Intercept)* | -8.35 | 0.03 | -8.78 | 0.07 |
| *age_category30-39* | 0.20 | 0.01 | 0.05 | 0.04 |
| *age_category40-49* | 0.07 | 0.01 | -0.03 | 0.04 |
| *age_category50-59* | -0.18 | 0.01 | -0.17 | 0.04 |
| *age_category60-69* | -0.35 | 0.01 | -0.33 | 0.04 |
| *age_category70-79* | -0.51 | 0.02 | -0.51 | 0.04 |
| *age_category80+* | -0.51 | 0.02 | -0.32 | 0.05 |
| *gendermale* | -0.17 | 0.01 | 0.09 | 0.02 |
| *vac_periodFebA* | -0.09 | 0.01 | -0.07 | 0.02 |
| *vac_periodFebB* | -0.19 | 0.01 | -0.07 | 0.03 |
| *vac_periodMarA* | -0.27 | 0.01 | -0.22 | 0.03 |
| *vac_periodMarB* | -0.34 | 0.01 | -0.26 | 0.04 |
| *vac_periodAprA* | -0.47 | 0.02 | -0.36 | 0.12 |
| *vac_periodAprB* | -0.61 | 0.04 | -1.70 | 0.71 |
| *vac_periodMayA* | -0.70 | 0.16 | -10.80 | 237.28 |
| *sectorgeneral_agas* | 0.31 | 0.01 | 0.13 | 0.03 |
| *sectororthodox_agas* | 0.70 | 0.01 | 0.64 | 0.04 |
| *incidence_group(2.34,3.33]* | 0.63 | 0.03 | 0.54 | 0.07 |
| *incidence_group(3.33,4.35]* | 0.87 | 0.03 | 0.84 | 0.06 |
| *incidence_group(4.35,5.36]* | 1.02 | 0.03 | 1.05 | 0.06 |
| *incidence_group(5.36,6.42]* | 1.22 | 0.03 | 1.15 | 0.06 |
| *incidence_group(6.42,7.51]* | *1.38* | *0.03* | *1.31* | *0.06* |
| *incidence_group(7.51,8.88]* | *1.51* | *0.03* | *1.38* | *0.06* |
| *incidence_group(8.88,10.5]* | *1.63* | *0.03* | *1.49* | *0.06* |
| *incidence_group(10.5,12.5]* | *1.77* | *0.03* | *1.60* | *0.06* |
| *incidence_group(12.5+]* | *2.05* | *0.02* | *1.92* | *0.06* |
| *pos_age_group60+:cohortbooster* | *-2.52* | *0.02* | *-2.00* | *0.03* |
| *pos_age_group50-59:cohortbooster* | *-2.50* | *0.03* | *-1.99* | *0.04* |
| *pos_age_group40-49:cohortbooster* | *-2.28* | *0.03* | *-1.69* | *0.04* |
| *pos_age_group30-39:cohortbooster* | *-2.18* | *0.04* | *-1.56* | *0.05* |
| *pos_age_group16-29:cohortbooster* | *-2.87* | *0.06* | *-2.42* | *0.07* |

**Table S9.** Poisson regression results for severe COVID-19.

| *term* | *Primary analysis* |  | *Secondary analysis* |  |
| --- | --- | --- | --- | --- |
|  | *estimate* | *std.error* | *estimate* | *std.error* |
| *(Intercept)* | -13.75 | 0.21 | -14.95 | 0.67 |
| *age_category50-59* | 1.08 | 0.17 | 0.07 | 0.56 |
| *age_category60-69* | 1.99 | 0.16 | 2.25 | 0.53 |
| *age_category70-79* | 2.80 | 0.16 | 3.07 | 0.52 |
| *age_category80+* | 3.74 | 0.16 | 4.21 | 0.51 |
| *gendermale* | 0.68 | 0.06 | 0.70 | 0.12 |
| *vac_periodFebA* | -0.17 | 0.07 | -0.03 | 0.14 |
| *vac_periodFebB* | -0.32 | 0.11 | 0.36 | 0.25 |
| *vac_periodMarA* | -0.46 | 0.11 | 0.05 | 0.35 |
| *vac_periodMarB* | -0.68 | 0.13 | 0.28 | 0.46 |
| *vac_periodAprA* | -0.64 | 0.23 | -13.50 | 759.93 |
| *vac_periodAprB* | -0.93 | 0.71 | -11.16 | 869.53 |
| *vac_periodMayA* | -6.92 | 326.10 |  |  |
| *sectorgeneral_agas* | -0.46 | 0.09 | -0.11 | 0.26 |
| *sectororthodox_agas* | -0.10 | 0.15 | -0.04 | 0.38 |
| *incidence_group(2.34,3.33]* | 0.54 | 0.17 | -0.08 | 0.52 |
| *incidence_group(3.33,4.35]* | 1.21 | 0.15 | 0.89 | 0.40 |
| *incidence_group(4.35,5.36]* | 1.21 | 0.16 | 0.61 | 0.40 |
| *incidence_group(5.36,6.42]* | 1.00 | 0.17 | 0.40 | 0.41 |
| *incidence_group(6.42,7.51]* | 1.32 | 0.16 | 0.80 | 0.39 |
| *incidence_group(7.51,8.88]* | 1.58 | 0.16 | 0.96 | 0.39 |
| *incidence_group(8.88,10.5]* | *1.68* | *0.15* | *0.77* | *0.40* |
| *incidence_group(10.5,12.5]* | *1.88* | *0.15* | *1.10* | *0.39* |
| *incidence_group(12.5+]* | *1.86* | *0.15* | *1.40* | *0.39* |
| *sev_age_group60+:cohortbooster* | *-2.93* | *0.09* | *-1.88* | *0.12* |
| *sev_age_group40-59:cohortbooster* | *-3.09* | *0.39* | *-1.16* | *0.56* |

**Table S10.** Poisson regression results for death due to COVID-19.

| *term* | *Primary analysis* |  | *Secondary analysis* |  |
| --- | --- | --- | --- | --- |
|  | *estimate* | *std.error* | *estimate* | *std.error* |
| *(Intercept)* | -13.21 | 0.32 | -13.99 | 0.94 |
| *age_category70-79* | 1.13 | 0.19 | 1.30 | 0.41 |
| *age_category80+* | 2.47 | 0.17 | 2.36 | 0.39 |
| *gendermale* | 0.87 | 0.12 | 0.85 | 0.26 |
| *vac_periodFebA* | -0.22 | 0.14 | -0.44 | 0.31 |
| *vac_periodFebB* | -0.06 | 0.22 | 0.25 | 0.60 |
| *vac_periodMarA* | -0.24 | 0.24 | -0.37 | 1.02 |
| *vac_periodMarB* | -0.76 | 0.40 | -13.44 | 824.72 |
| *vac_periodAprA* | -13.17 | 836.00 |  |  |
| *sectorgeneral_agas* | -0.77 | 0.21 | -0.21 | 0.62 |
| *sectororthodox_agas* | -0.31 | 0.33 | -0.53 | 0.92 |
| *incidence_group(2.34,3.33]* | 0.36 | 0.32 | 0.30 | 1.00 |
| *incidence_group(3.33,4.35]* | 1.50 | 0.27 | 1.41 | 0.78 |
| *incidence_group(4.35,5.36]* | 1.43 | 0.29 | 1.06 | 0.79 |
| *incidence_group(5.36,6.42]* | 0.91 | 0.35 | 0.40 | 0.87 |
| *incidence_group(6.42,7.51]* | 1.49 | 0.31 | 0.79 | 0.80 |
| *incidence_group(7.51,8.88]* | 1.67 | 0.30 | 1.02 | 0.78 |
| *incidence_group(8.88,10.5]* | 2.06 | 0.29 | 0.35 | 0.85 |
| *incidence_group(10.5,12.5]* | 1.99 | 0.32 | 0.72 | 0.84 |
| *incidence_group(12.5+]* | 2.06 | 0.32 | 1.66 | 0.79 |
| *cohortbooster* | -2.69 | 0.23 | -1.57 | 0.27 |

**Table S11.**
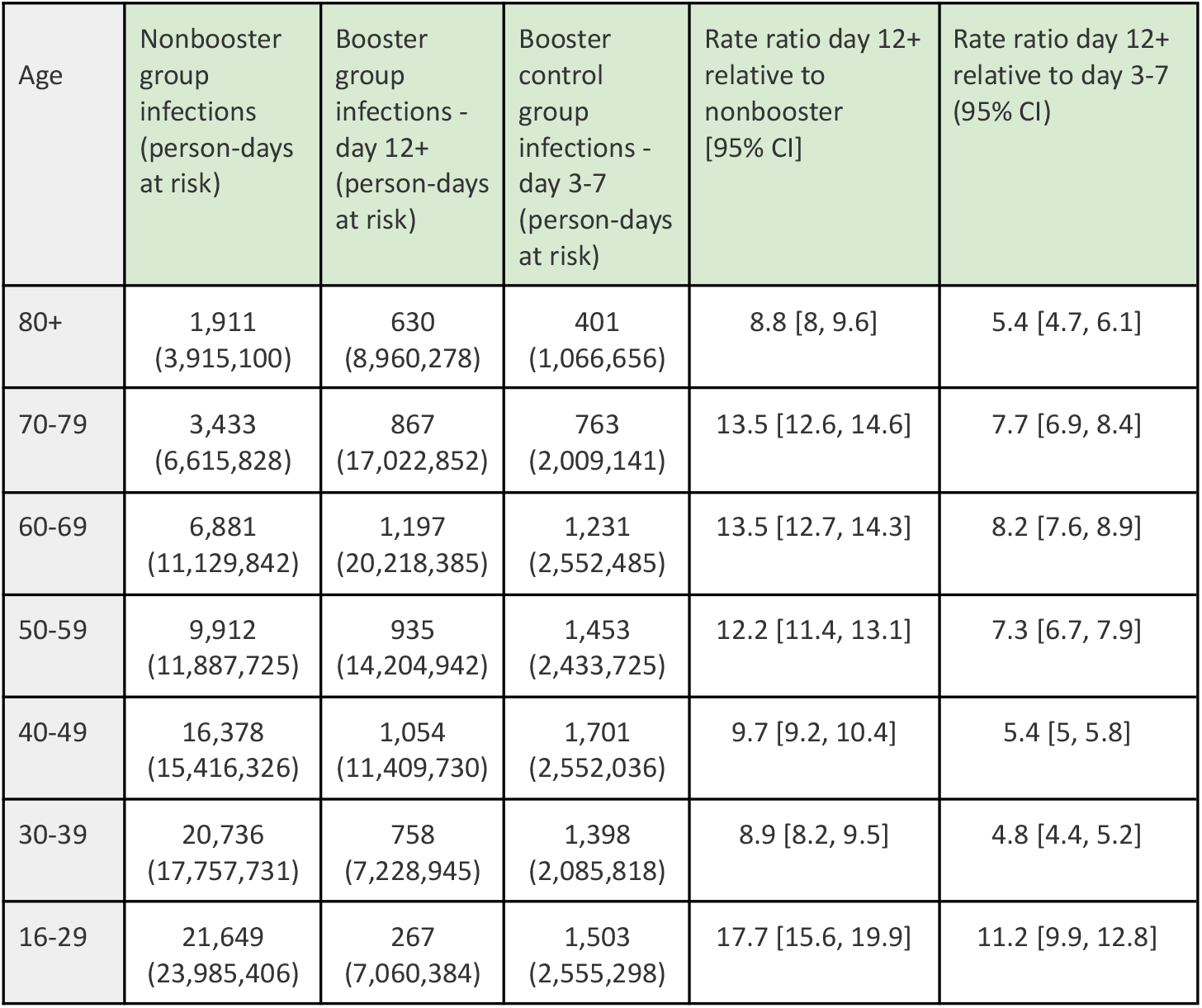
Summary of the results regarding confirmed infections of the Poisson regression analysis for the booster group and nonbooster group, with breakdown for age groups above 60 years of age. For each group, we provide the number of confirmed infections, the total number of person-days at risk, and the estimated rate ratio for the primary analysis (nonbooster relative to at least 12 days post booster-vaccination) and the secondary analysis (3-7 days post booster-vaccination relative to at least 12 days post booster-vaccination).

| Age | Nonbooster group infections (person-days at risk) | Booster group infections - day 12+ (person-days at risk) | Booster control group infections - day 3-7 (person-days at risk) | Rate ratio day 12+ relative to nonbooster [95% CI] | Rate ratio day 12+ relative to day 3-7 (95% CI) |
| --- | --- | --- | --- | --- | --- |
| 80+ | 1,911<br>(3,915,100) | 630<br>(8,960,278) | 401<br>(1,066,656) | 8.8 [8, 9.6] | 5.4 [4.7, 6.1] |
| 70-79 | 3,433<br>(6,615,828) | 867<br>(17,022,852) | 763<br>(2,009,141) | 13.5 [12.6, 14.6] | 7.7 [6.9, 8.4] |
| 60-69 | 6,881<br>(11,129,842) | 1,197<br>(20,218,385) | 1,231<br>(2,552,485) | 13.5 [12.7, 14.3] | 8.2 [7.6, 8.9] |
| 50-59 | 9,912<br>(11,887,725) | 935<br>(14,204,942) | 1,453<br>(2,433,725) | 12.2 [11.4, 13.1] | 7.3 [6.7, 7.9] |
| 40-49 | 16,378<br>(15,416,326) | 1,054<br>(11,409,730) | 1,701<br>(2,552,036) | 9.7 [9.2, 10.4] | 5.4 [5, 5.8] |
| 30-39 | 20,736<br>(17,757,731) | 758<br>(7,228,945) | 1,398<br>(2,085,818) | 8.9 [8.2, 9.5] | 4.8 [4.4, 5.2] |
| 16-29 | 21,649<br>(23,985,406) | 267<br>(7,060,384) | 1,503<br>(2,555,298) | 17.7 [15.6, 19.9] | 11.2 [9.9, 12.8] |

**Table S12.** Summary of the results regarding severe COVID-19 cases of the Poisson regression analysis for the booster group and nonbooster group, with breakdown for age groups above 60 years of age. For each group, we provide the number of confirmed infections, the total number of person-days at risk, and the estimated rate ratio for the primary analysis (nonbooster relative to at least 12 days post booster-vaccination) and the secondary analysis (3-7 days post booster-vaccination relative to at least 12 days post booster-vaccination).

| Age | Nonbooster severe cases (person-days at risk) | Booster group severe cases - day 12+ (person-days at risk) | Booster control group severe cases - day 3-7 (person-days at risk) | Rate ratio day 12+ relative to nonbooster [95% CI] | Rate ratio day 12+ relative to day 3-7 (95% CI) |
| --- | --- | --- | --- | --- | --- |
| 80+ | 424<br>(3,774,512) | 80<br>(7,706,159) | 60<br>(1,056,597) | 15.2 [11.9, 19.4] | 5.8 [4.1, 8.1] |
| 70-79 | 306<br>(6,408,860) | 41<br>(14,659,120) | 47<br>(1,988,328) | 24.9 [17.9, 34.6] | 9.1 [6, 13.9] |
| 60-60 | 227<br>(10,711,374) | 29<br>(17,264,761) | 20<br>(2,503,853) | 17.7 [12, 26.2] | 5 [2.8, 8.9] |

## Notes

### Competing Interest Statement

The authors have declared no competing interest.

### Funding Statement

None

### Author Declarations

The study was approved by the institutional review board of the Sheba Medical Center (Helsinki approval number: SMC-8228-21).

